# Estimating the contagiousness ratio between two viral strains

**DOI:** 10.1101/2023.04.27.23289192

**Authors:** Giulia Della Croce Di Dojola, Gianluca Mastrantonio, Francesco Cerutti, Valeria Ghisetti, Mauro Gasparini, Enrico Bibbona

## Abstract

We propose a new method to estimate the ratio between the basic reproduction numbers of a newly emerged variant and the one currently dominating. We use public data of two kinds: the proportions of the daily infected from each strain obtained from a random sample that has been sequenced, and the epidemic curves of total infections and recoveries. Our method is based on a new discrete-time SIR model with two strains, considered both in the deterministic and stochastic versions. In the deterministic case we use maximum likelihood, while in the stochastic setting, since we need to reconstruct the missing incidence and prevalence information of the new variant, we decided to use a hierarchical Bayesian hidden Markov model. This new methodology is applied to data from the Piedmont Italian region in December-January 2022, when the Omicron variant started to be observed and quickly became prevalent. With both approaches, we obtain an estimate of the contagiousness ratio that is consistent with other studies specifically designed for the aim.

## 1. Introduction

Natural selection has shaped the evolution of all living beings, including viruses. Differently from other organisms, the life-cycle of a viral particle is very short (hours or days) and the time scale of the evolution is such that a single generation of humans can witness the flow of thousands of virus generations, the appearance of several mutations and the effect of natural selection. This mechanism has been particularly visible during the recent COVID-19 pandemic, in which several variants have replaced one another under the selective pressure of the human immune system. Whenever a new variant emerges, one of the targets of the surveillance policies that are typically undertaken is the early evaluation of its characteristics, including its ability to transmit (Lyngse *and others* (2022); Campbell *and others* (2021)).

Several articles have been published assessing a comparison of the contagiousness of the different variants, the Alpha variant with respect to the preexisting lineages (Volz *and others* (2021); Davies *and others* (2021)), the Delta variant with respect to the ancestral strain (Liu and Rocklöv (2021)) or a comparison of the effective reproduction number of the different observed strains (Campbell *and others* (2021); Bhatia *and others* (2021)), all strains up to Delta, and finally the Omicron variant versus Delta (Liu and Rocklöv (2022); Ito *and others* (2022)).

The methodology used for these evaluations is very heterogeneous, and the datasets used are themselves of different kinds. The aim of this paper is to introduce a statistical methodology to estimate the ratio of the effective reproduction numbers of a newly emerged strain with respect to the one currently dominating, without resorting to external information such as an estimate of the basic reproduction number of the latter, and without fixing the value of any parameter. The data on which we base our analysis are the epidemic curves, which are publicly available for many countries in easily accessible repositories, and the proportions of sequenced samples that are attributed to the new variant, again publicly available for several nations. The method we propose, moreover, is based on new mathematical models which generalise well-established counterparts to a virus with two strains such that, an individual who recovers from any of the two infections, acquires immunity to both.

## 2. Data

Consider an infectious disease transmitted by a virus with two strains, with mutual immunity, that compete for the infection of the population. Assume that at the beginning of the analysis strain 1 is dominating, while strain 2 is emerging during the time window of the study. For each day *j*, we assume the data consist in:

- *n*_*j*_ samples randomly selected from the new daily infections and sent to sequencing facilities;
- *z*_*j*_ newly sequenced samples attributed to strain 2 (being *z*_*j*_*/n*_*j*_ a proxy for its relative incidence). Data of this kind have been made publicly available from different national authorities during the COVID-19 pandemic (e.g. Istituto Superiore di Sanitá (2023), Staten Serum Institut (2022), UK Health Security Agency (2021)). Furthermore, the epidemic curves of daily new infections and daily active infections are needed for each *j*. Such data are also publicly available (Presidenza del Consiglio dei Ministri - Dipartimento della Protezione Civile (2023)) and here they are indicated as:
- 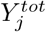 total new cases (a proxy for the incidence), including both strain 1 and strain 2;
- 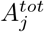 total active cases (a proxy for the prevalence), including both strain 1 and strain 2.

From these quantities, the new recoveries 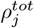 of each day can be directly derived

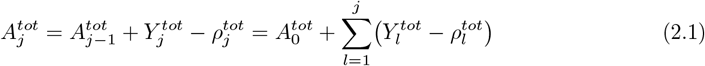

Vice versa, if the 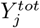 and 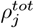 are known for each day, the 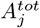 can be derived with the same relation, provided that 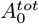 is known.

Let us remark that 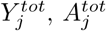, and 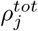 could be in principle split according to 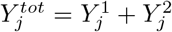, where 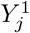 is the number of new cases due to strain 1, and similarly 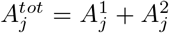, and 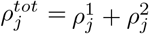. However, 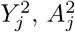, and 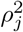 are most often not directly observed, which is the reason why they are here considered as latent variables, to be estimated in our inferential procedure.

## 3. Mathematical model

To define what we mean by contagiousness ratio and how we use our data to estimate it (latent variables included) we need to introduce a mathematical model of the epidemic spread. Since our data are daily observations, discrete-time models are adopted. In particular, two different discrete-time models are estimated, one deterministic and one stochastic. To better explain the reasoning and the features of each model, we first introduce them in their continuous-time version, and then we use a discretisation procedure to derive the final model. All models we propose here can be seen as a compartmentalisation model, whose general structure is depicted in Figure 1.

**Fig. 1.**
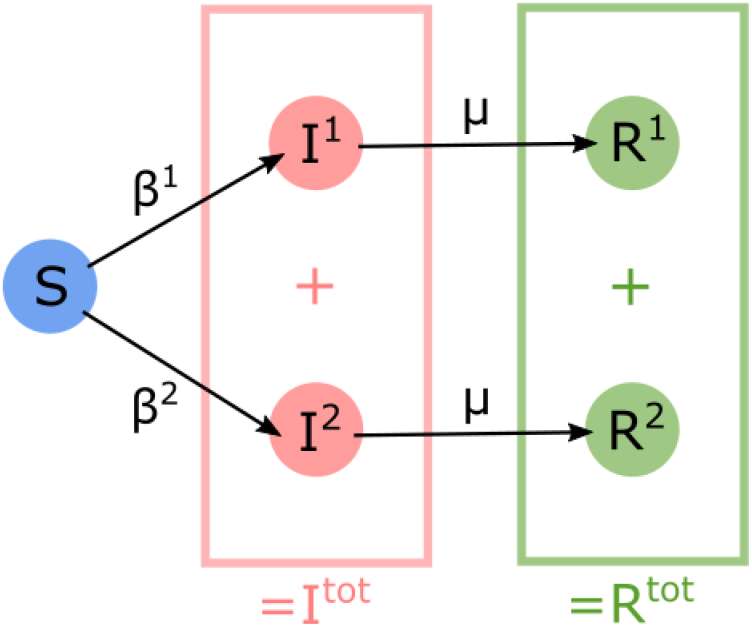
Compartments of the two strains epidemic models introduced in the paper.

### 3.1 Deterministic models

The best-known deterministic epidemic model is the SIR model, established in Kermack and McKendrick (1927). In this section, we generalise it to the case of a viral infection that is caused by two strains with mutual immunity, and we introduce a suitable discrete-time version.

#### 3.1.1 Continuous-time

For each time *t* ⩾ 0, let *s*(*t*) denote the population of susceptible individuals, *i*^*v*^(*t*) the population of individuals infected by strain *v* (*v* ∈ {1, 2}), and *r*^*v*^(*t*) the population of individuals recovered from strain *v*. The size of the total population is *N*. A natural extension of the SIR model is provided by the following system of ordinary differential equations (ODE):

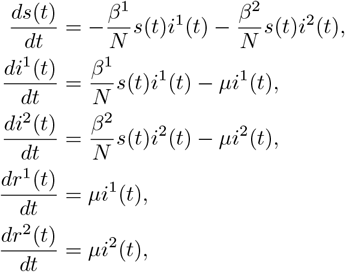

where parameters *β*^1^ and *β*^2^ can be interpreted as rates of infection, for strain 1 and 2 respectively, while *μ* is the recovery rate, that is assumed to be the same for the two variants. Hence, the two basic reproduction numbers are equal to 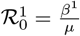 and 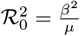.

We define the following parameter:

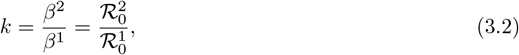

that describes how many times one variant is more contagious than the other one, and it also coincides with the ratio between the basic reproduction numbers (ℛ_0_). Estimating this parameter (and its discrete-time stochastic/deterministic equivalent) is the target of our analysis.

#### 3.1.2 Discrete-time

A discrete-time version of the SIR model previously introduced is obtained by the Euler discretisation method, with stepsize Δ*t*. For every step *j*, we have the subpopulations 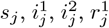 and 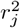, with 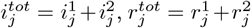 and 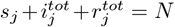. The model equations are:

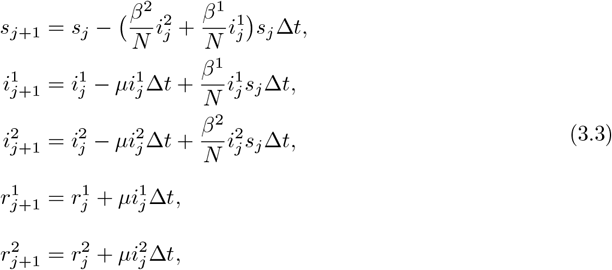

where Δ*t* is the time interval between two consecutive steps. This model and the continuous one in Section 3.1.1 converge when Δ*t* tends to 0. In our application we use (3.3) to model daily data (Δ*t* = 1). In this regime, the agreement between the two models depends on the values of the parameters.

Parameter *k*, defined as in Equation (3.2), is again interpreted as a *contagiousness ratio*. The model presented in this section is not well-defined, since states are allowed to be negative. For our analysis, this problem does not have any practical consequence (real data are positive), but if used for other purposes it may be necessary to modify it in the neighbourhood of the boundaries of the positive orthant.

### 3.2 Stochastic models

We introduce a stochastic SIR model with two strains, first in continuous-time, and then in discrete-time.

#### 3.2.1 Continuous-time

The state of the system at time *t* is expressed by a positive integer vector **X**(**t**) = (*S*(*t*), *I*^1^(*t*), *I*^2^(*t*), *R*^1^(*t*), *R*^2^(*t*))′, describing how many individuals are in each compartment. This notation is redundant since, ∀*t, S*(*t*) + *I*^1^(*t*) + *I*^2^(*t*) + *R*^1^(*t*) + *R*^2^(*t*) = *N*. At each time instant *t* ⩾ 0, the allowed transitions are four: infection and recovery for each of the two variants. The corresponding rates at a general state (*s, i*^1^, *i*^2^, *r*^1^, *r*^2^) are

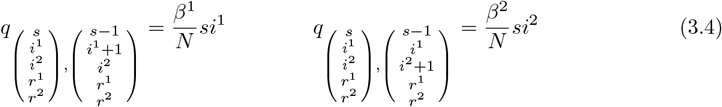

for infections, and

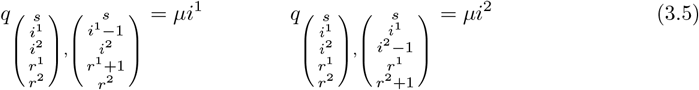

for recoveries.

Due to a well-known result on the fluid limit of density-dependent Markov chains (Kurtz (1970)), the continuous-time stochastic model just described converges to the deterministic one in Section 3.1.1 as *N* tends to infinity.

#### 3.2.2 Discrete-time

The discrete analogue of the previous model is based on the Binomial Tau-Leap discretisation method (Chatterjee *and others* (2005)), with stepsize Δ*t*. It is a two-strain generalisation of Scalia Tomba (1990), which, in turn, generalises the well-known Reed Frost model. Given the state 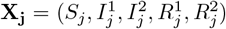, the system is described by the following dynamics

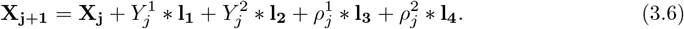

In the previous equation, **l_1_, l_2_, l_3_**, and **l**_**4**_ are the vectors that represent the state increments caused by each transition. Such vectors are: **l_1_** = (−1, +1, 0, 0, 0)′ for the infection by strain 1, **l_2_** = (−1, 0, +1, 0, 0)′ for the infection by strain 2, **l_3_** = (0, −1, 0, +1, 0)′ for the recovery from strain 1 and **l**_**4**_ = (0, 0, −1, 0, +1)′ for the recovery from strain 2. The terms 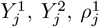 and 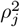 represent, for each step *j*, the new infections by strain 1 and strain 2 and the new recoveries from strain 1 and strain 2. They are distributed according to the following laws

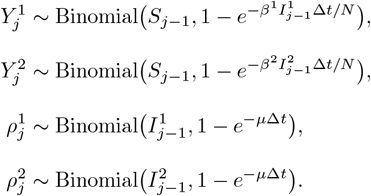

This model and the continuous one in Section 3.2.1 converge when Δ*t* tends to 0. Our application uses (3.6) to model daily data (Δ*t* = 1). Again, in this regime, the agreement between the two models depends on the values of the parameters.

## 4. Statistical model and method

The goal of this Section is to define a statistical model and a methodology to estimate the contagiousness ratio *k* of Equation (3.2), assuming that the epidemics follows a discrete-time model, which is either the deterministic one in Section 3.1.2 or the stochastic one in Section 3.2.2. Based on the available data (cf. Section 2), at each day *j, n*_*j*_ samples are extracted from the new infections 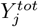 and sent to the sequencing facilities. Among those, *z*_*j*_ are attributed to strain 2. For the days in which sequencing is not performed, we consider *n*_*j*_ = *z*_*j*_ = 0. If the counts 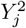 of new infections due to strain 2 were known, it would be natural to state that *z*_*j*_ follows the hypergeometric distribution

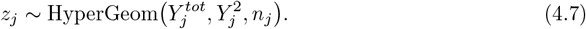

However, the 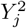 are latent quantities, since the total counts only are given as data, but we don’t know how many are caused by each variant. We aim to reconstruct the missing information through the discrete-time models introduced in Sections 3.1.2 and 3.2.2.

For the latent strain 2 quantities 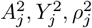 the following recursive relation holds

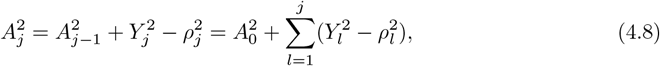

given an initial value 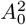 that is considered an additional model parameter. Equation (4.8) is the analogue of Equation (2.1) for the corresponding total quantities, and a similar recurrence relation holds for 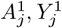, and 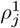.

### 4.1 Deterministic setting

Based on the deterministic discrete-time SIR model with daily observations (Δ*t* = 1), we assume that the data and the latent quantities are related to the model equations in Formula (3.3) by

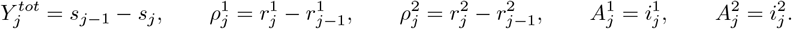

The latent variables 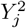 and 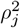 can be reconstructed using the deterministic relationships that we now introduce. We have

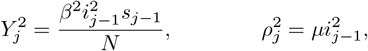

which can be recast in the following form

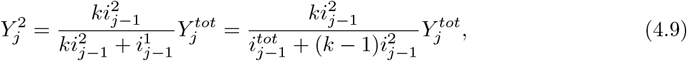

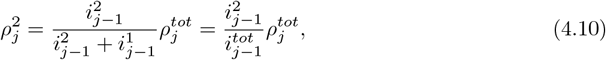

which depends on the contagiousness ratio *k*, cf. Equation (3.2), the known total quantities 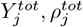, and the latent 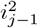.

Concerning the 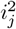 latent quantities, they can be recursively computed substituting the Equations (4.9) and (4.10) in Equation (4.8), obtaining

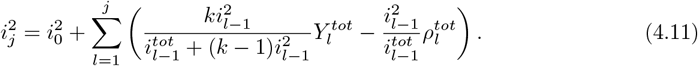

Therefore, each 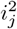 is a function of the data and of the parameters 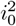 and *k* only. Substituting back the expression for 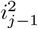 in Equation (4.9), the latent quantities 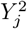 are now completely determined. The log-likelihood of the *z*_*j*_ can be explicitly computed and results in a function of the data and model parameters only

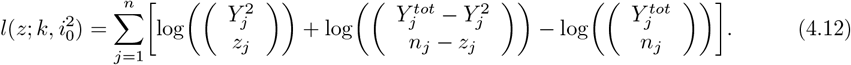

Maximisation of this quantity provides maximum likelihood (ML) point estimates for *k* and 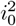. Confidence intervals can be derived both using the profile likelihood and by parametric bootstrap. It is important to notice that the binomial coefficients above are meant in their generalized form, based on the Euler gamma function, so that the argument 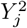 needs not be an integer. Some care in the calculation of the likelihood is needed since the values of *k* and 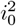 that lead to 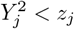 or 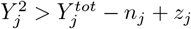 are not acceptable, and they should be forced to have zero likelihood.

### 4.2 Stochastic setting

Based on the stochastic discrete-time SIR model with daily data (Δ*t* = 1), we assume that the data and the latent quantities are related to the model equations in Formula (3.6) by

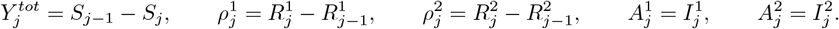

In this setting, 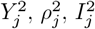 are random quantities that need to be inferred. To this aim, we adopt a Bayesian framework, based on an Hidden Markov Model (HMM). The hidden process is given by the couple 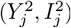, *j* = 1, …, *n*, while the observed process consists in the *z*_*j*_ variables. Along with the hidden process, the parameters *k* and 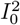 are the target of our inference. The resulting hierarchical model is, for all *j* = 1, …, *n*,

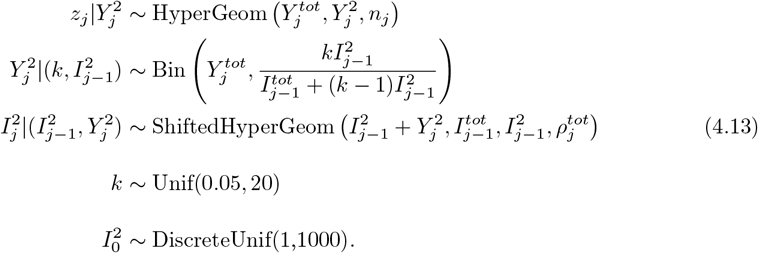

The shifted hypergeometric conditional distribution of 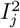, denoted as ShiftedHyperGeom above, is defined as follows: *A* is ShiftedHyperGeom, i.e., *A* ∼ ShiftedHyperGeom(*a, b, c, d*), if and only if ℙ (*A* = *i*) = ℙ (*B* = *a* − *i*), where *B* ∼ HyperGeom(*b, c, d*). This choice is motivated by Equation (4.8) and the assumption that for all *j* = 1, …, *n*

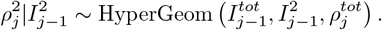

The conditional log-likelihood of *z*_*j*_, given 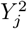, has the same expression as in the deterministic case (Equation (4.12)), but now we reinterpret 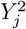 as a random variable. The model dependencies are illustrated in Figure 2.

**Fig. 2.**
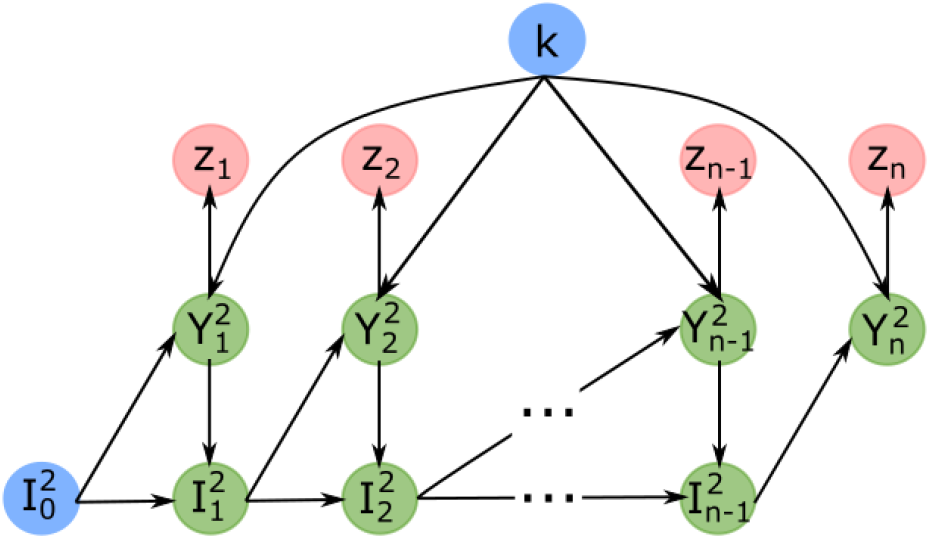
Graphical representation of the stochastic model defined in the paper.

Our inferential procedure is based on an MCMC algorithm that targets the posterior distributions of the parameters *k* and 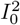, according to the hierarchical model just explained. We sample *k*, 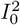and the hidden trajectories 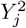 and 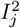 by a standard Metropolis within Gibbs algorithm, whose details are described below.

#### 4.2.1 Proposal distributions

The proposal distribution used for parameter *k* is a normal distribution on *τ* = log(*k*), with mean equal to the logarithm of the current value of *k* and with a standard deviation of 0.02. For the discrete terms 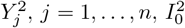, and 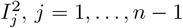, *j* = 1, …, *n* − 1, we use a discrete uniform distribution around the current value of the chain. To specify the ranges of these uniform distributions, some admissibility constraints need to be imposed. For any given day *j* in {1, …, *n* − 1}, we need to fulfil the four easily interpretable conditions listed below.

Interpretable condition Same condition in terms of the hidden state

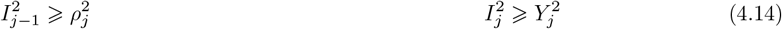

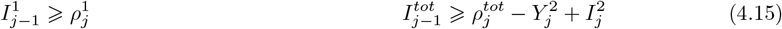

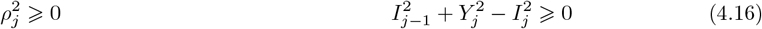

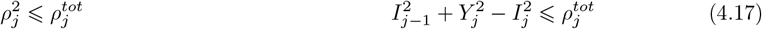

In order to turn such conditions in terms of the state variables 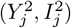, we apply Equation (4.8) and rewrite the strain 1 quantities as the difference between the total quantities and the strain 2 quantities. For each variable being proposed, we have to impose the four constraints above, together with other natural bounds, that are introduced case by case.

##### Updating 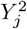

Let us denote with 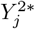 the proposed value, and with 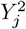 the current value of the chain. Any proposed value 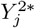, in order to be compatible with the sequencing data, has to satisfy the following

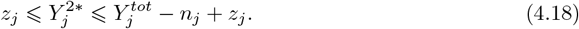

Furthermore, the constraints in Equations (4.14), (4.15), (4.16) and (4.17), expressed in terms of 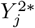, are

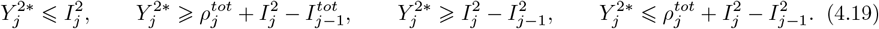

Finally, by summarizing Equations (4.18) and (4.19), the bounds for the uniform proposal of 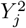, for all *j* = 1, …, *n* − 1, are

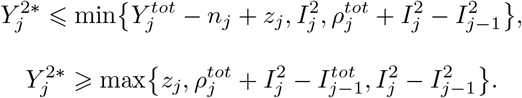

For the update of 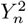, we only need to impose Equation (4.18), with *j* = *n*.

##### Updating 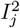

When proposing a new value 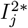 (with 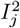 denoting the current value of the chain), the constraints in Equations (4.16) and (4.17) have to be considered not only with respect to 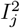, but also with respect to 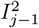, giving rise to the following bounds

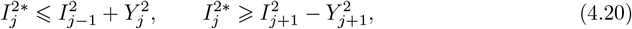

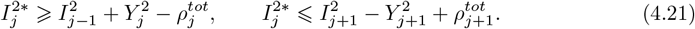

Equations (4.14) and (4.15) are simply rewritten in terms of 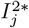

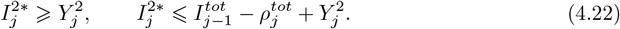

Summarising all the previous constraints, together with 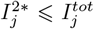 to maintain agreement with the epidemic data, the final bounds for the uniform proposal of 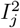, for all *j* = 1, …, *n* − 2, are

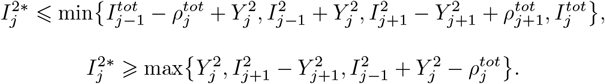

For 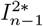 only the first constraint in Equation (4.20), the first constraint in Equation (4.21), and both constraints in Equation (4.22), together with 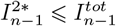, have to be imposed. All have to be meant with *j* = *n* − 1. At last, the bounds for the uniform proposal of 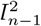 are

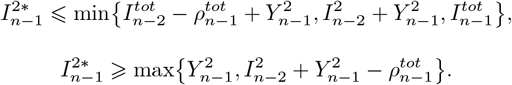

##### Updating 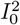

When proposing a new value 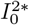 (being 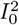 the current value in the chain), we impose the second constraint of Equation (4.20) and the second constraint of Equation (4.21) (both considered with *j* = 0), together with 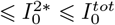. The final bounds for the uniform proposal of 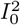 are

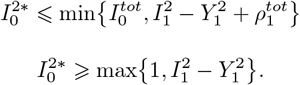

## 5. Results

The methods presented in the previous section are applied first to simulated data, to ensure the validity of the models, to check their computational cost, and the correctness of the implementations. Then, we address publicly available data on the Delta (strain 1) and Omicron (strain 2) variants of the SARS-CoV-2 virus, in the Italian region Piemonte, with a population size of about *N* = 5 · 10^6^. This specific choice is motivated by the research project SORGENTE (SORveglianza GENomica in PiemonTE), funded by the Piemonte Region, in which our team participates. The epidemic curves, consisting of total counts of daily active cases and new infections, can be found at Presidenza del Consiglio dei Ministri - Dipartimento della Protezione Civile (2023), while sequencing data are from the Quick Surveys conducted by the *ISS* (*Istituto Superiore di Sanitá*). We analyse the period in which the Omicron variant started to be observed and quickly became prevalent. In this period, the Quick Surveys have been carried out every two weeks, resulting in five data points; data are shown in Table 1.

**Table 1.**
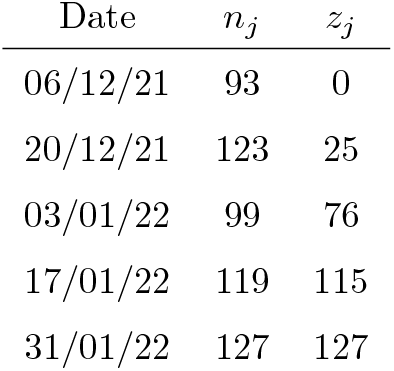
*ISS* sequencing data (Istituto Superiore di Sanitá (2023)).

### 5.1 Simulated data

Both the deterministic and the stochastic approach have been tested on different synthetic datasets. A detailed account of the results is presented in the Supplementary Material, available online. The main conclusions are that both methods consistently estimate the model parameters, and recover a relative incidence curve with a good fit of the data. The Bayesian approach is computationally expensive, since, at each iteration, the value of the hidden process can only be updated separately at each day. Indeed, the forward-backward approach proposed by Chib (1996) to update the entire trajectory at once is unfeasible, due to the huge size of the state space that does not allow to store all the entries of the transition matrix. This causes slow mixing and a strong correlation of the chains, which need to be thinned heavily.

### 5.2 Real data, results with the deterministic approach

To obtain the parameter estimates, we maximise the log-likelihood function in Equation (4.12) using the `mle` R function, with the `L-BFGS-B` method (R Core Team, 2022). We obtain the estimates shown in Table 2, together with the 95% confidence intervals, which are computed both using the profile likelihood and by applying the parametric bootstrap method with a Monte Carlo simulation of 1000 synthetic samples.

**Table 2.** ML estimates and 95% confidence intervals of the parameters, deterministic approach.

| Parameter | Estimate | 95% profile C. I. | 95% C.I. parametric bootstrap |
| --- | --- | --- | --- |
| $k$ | 4.4 | [3.7, 5.48] | [3.81, 5.64] |
| $i_0^2$ | 21.3 | [5.73, 64.51] | [4.52, 55.4] |

Table 2 shows that the Omicron variant is 4 to 5 times more contagious than Delta and that at the beginning of our analysis, 5 December 2021, despite Omicron was still not detected, there were around 20 Omicron cases already circulating within the Piedmont region. Using these values, it is possible to reconstruct the estimated Omicron relative incidence curve, to be compared with data. The comparison is shown in Figure 3.

**Fig. 3.**
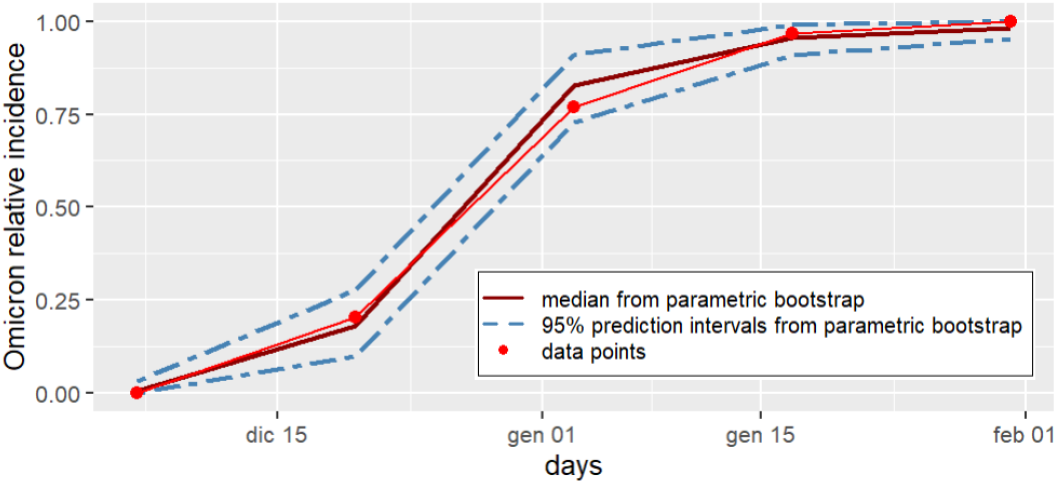
Estimated Omicron relative incidence curve against the data points in Table 1, deterministic approach.

### 5.3 Real data, results with the stochastic approach

We run the MCMC algorithm for 6 · 10^7^ iterations, with a burn-in of 3 · 10^6^ and thinning every 10^4^ runs. These values are chosen since, with extensive simulation study (cf. Section 5.1), they prove to give posterior samples that are independent and sampled from the posterior distribution. The trace plots and the related posterior distributions are shown in Figures 4 and 5. The coloured lines in the trace plots refer to the posterior medians, that are quantified in Table 3, together with the 95% credible intervals.

**Fig. 4.**
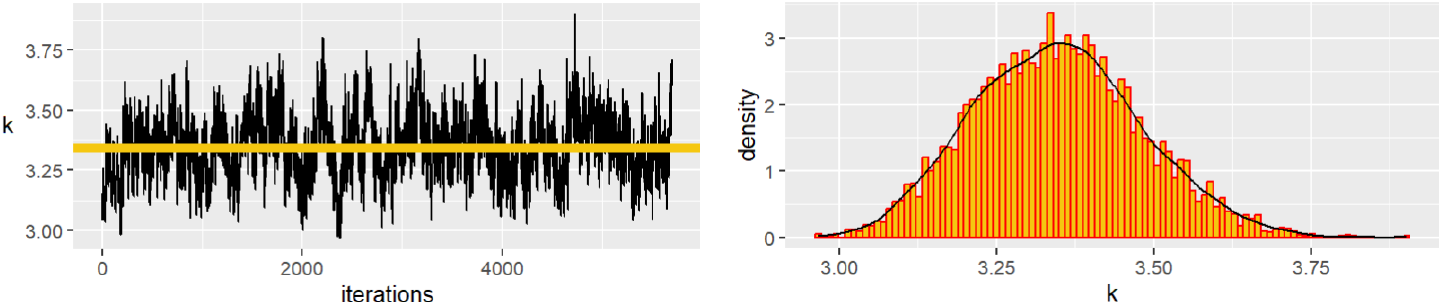
Trace plot and posterior distribution of parameter *k*, stochastic approach.

**Fig. 5.**
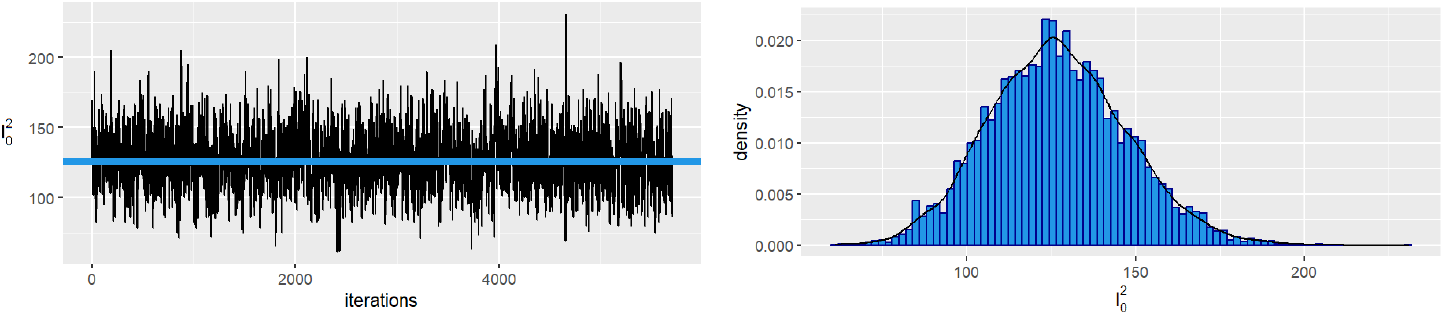
Trace plot and posterior distribution of parameter 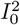, stochastic approach.

**Table 3.** Posterior median and 95% credibility intervals of the parameters, stochastic approach.

| Parameter | Posterior Median | 95% C.I. |
| --- | --- | --- |
| $k$ | 3.34 | [3.1, 3.6] |
| $I_0^2$ | 126 | [88, 169] |

Despite the chains being already heavily thinned, a non-negligible autocorrelation persists in the trace plot of *k*, even at lags as large as 60 (see Figure 6). This fact does not harm our analysis, which is based on 5700 posterior samples, and it is to be traced back to an expected inefficiency of the sampling algorithm which updates the hidden points 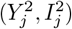 one by one, cf. Section 5.1.

**Fig. 6.**
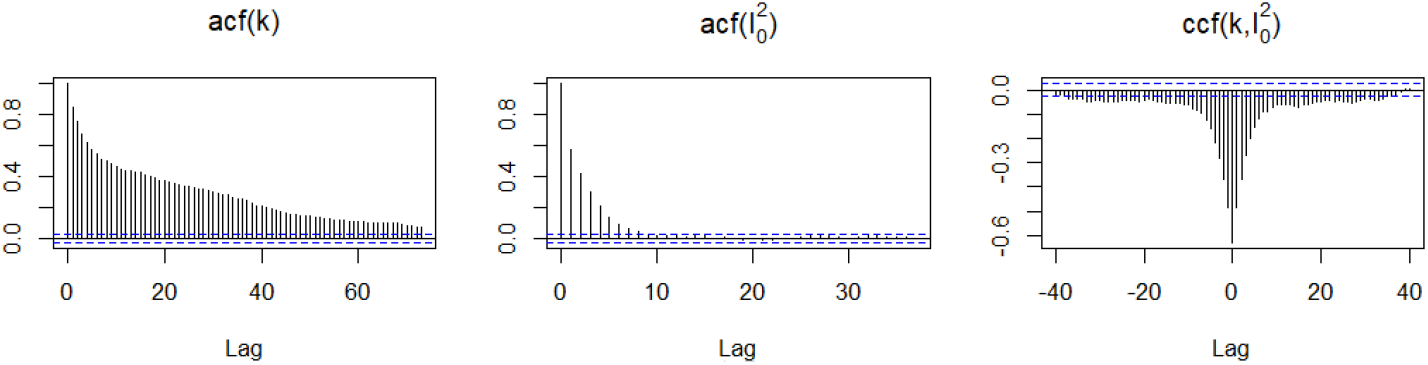
Autocorrelation and Cross-correlation of the parameters, stochastic approach.

In Figure 7 we plot the posterior median of the estimated Omicron relative incidence curve, together with the 95 percentiles of the predictive distribution. These are obtained from each posterior sample of the parameters, sampling from the conditional distribution of the *z*_*j*_ (first equation of (4.13)). Experimental data points are well captured and contained within such intervals.

**Fig. 7.**
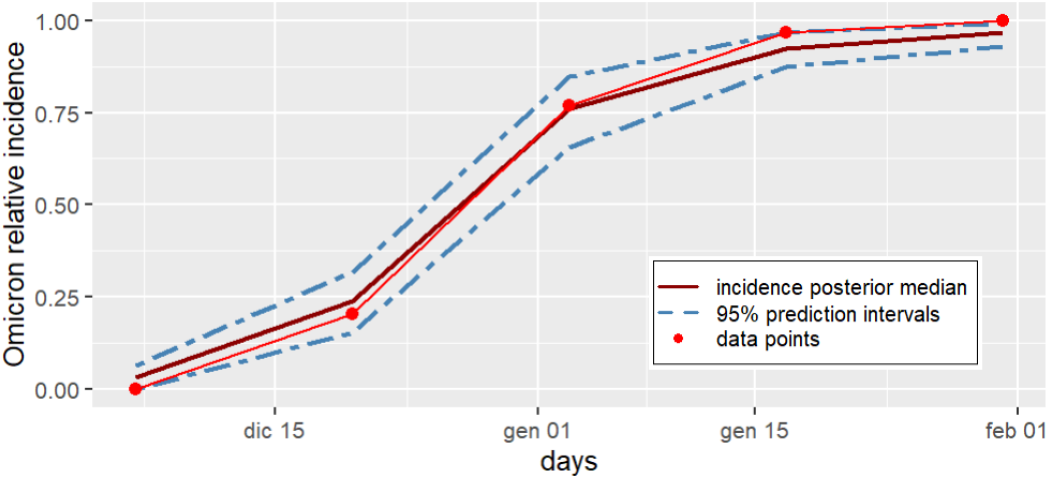
Posterior median and 95% prediction intervals for the estimated Omicron relative incidence curve, stochastic approach.

## 6. Discussion

We recall the main advantages of the methods here proposed. First of all, it can be applied to publicly available data, that have been collected in several countries and that are not subject to any privacy concerns. Second, we do not need to rely on any external estimation of the model parameters, such as the basic reproduction number of the pre-existing variant, since we directly target a ratio, that we can estimate using the available data alone. Third, our model is based on solid mathematical ground, without artificial assumptions.

To compare the deterministic and the stochastic approach, we observe that both Omicron relative incidence curves fit data properly, with the five data points contained within the prediction intervals. Compared to the prediction intervals obtained with the deterministic approach, the stochastic ones are very similar in the central days but larger in the initial and final phases. As for the estimated parameters, the stochastic model tends to prefer a smaller value of *k* and a higher 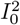, being the two negatively correlated (see Figure 6, third panel). The estimate of *k* from the stochastic model is in very good agreement with the estimates available in the literature from different methods, e.g. Ito *and others* (2022) and Liu and Rocklöv (2022), while the estimate obtained from the deterministic models is a bit higher. We believe that the stochastic model may be more robust in a context like ours, where the population is of an intermediate size. On the other hand, the computational cost is beyond compare, a few seconds for the ML deterministic approach, and a couple of days for the Bayesian stochastic one.

In the context of massive daily sequencing, such as what was implemented in Denmark and the UK (cf. Staten Serum Institut (2022); UK Health Security Agency (2021)), our method could be embedded into a surveillance system to give an early estimate of the transmission advantage of a newly emerged variant and potentially raise an alarm before it starts spreading to other regions. To this aim, it would be useful to evaluate how many days are needed to reach a sufficiently good estimate of *k* to determine whether or not the new variant deserves some concern.

## Data Availability

All data produced are available online at:
Istituto Superiore di Sanita', Monitoraggio delle varianti del virus SARS-CoV-2 di interesse in sanita' pubblica in Italia, https://www.epicentro.iss.it/coronavirus/sars-cov-2-monitoraggio-varianti-indagini-rapide
Presidenza del Consiglio dei Ministri - Dipartimento della Protezione Civile, https://github.com/pcm-dpc/COVID-19

https://github.com/pcm-dpc/COVID-19

## Software

The data and the R code used for the analysis are available at https://github.com/giuliadellacroce/sorgente.git.

## Author contribution

GDC and EB formulated the project, defined the epidemic model and the estimation methods, and drafted the manuscript. GDC wrote the code. GM gave guidance for the MCMC algorithm. EB, GM, MG, FC, and VG supervised the project and the writing of the manuscript.

## Supplementary Material

Supplementary material is available online.

## Acknowledgments

We acknowledge funding from Regione Piemonte, INFRA-P2 COVID, Reg. (UE) 1407/2013, CID 378-26, Progetto SORGENTE, CUP E15F21003680007. The authors are indebted to Greg Rempala, Ohio State University, for some fruitful discussions out of which this project was started.

## Conflict of Interest

None declared.

## Notes

### Competing Interest Statement

The authors have declared no competing interest.

### Author Declarations

Istituto Superiore di Sanita', Monitoraggio delle varianti del virus SARS-CoV-2 di interesse in sanita' pubblica in Italia, https://www.epicentro.iss.it/coronavirus/sars-cov-2-monitoraggio-varianti-indagini-rapide Presidenza del Consiglio dei Ministri - Dipartimento della Protezione Civile, https://github.com/pcm-dpc/COVID-19

## References

Bhatia, S., Wardle, J., Nash, R. K., Nouvellet, P. and Cori, A. (2021). A generic method and software to estimate the transmission advantage of pathogen variants in real-time: Sars-cov-2 as a case-study. medRxiv. doi:10.1101/2021.11.26.21266899.

Campbell, F., Archer, B., Laurenson-Schafer, H., Jinnai, Y., Konings, F., Batra, N., Pavlin, B., Vandemaele, K., Van Kerkhove, M. D., Jombart, T., Morgan, O. and others. (2021). Increased transmissibility and global spread of sars-cov-2 variants of concern as at june 2021. Eurosurveillance 26(24).

Chatterjee, A., Vlachos, D. G. and Katsoulakis, M. A. (2005). Binomial distribution based τ-leap accelerated stochastic simulation. The Journal of Chemical Physics 122(2), 024112.

Chib, S. (1996). Calculating posterior distributions and modal estimates in markov mixture models. Journal of Econometrics 75(1), 79–97.

Davies, N. G., Abbott, S., Barnard, R. C., Jarvis, C. I., Kucharski, A. J., Munday, J. D., Pearson, C. A. B., Russell, T. W., Tully, D. C., Washburne, A. D., Wenseleers, T., Gimma, A., Waites, W., Wong, K. L. M., van Zandvoort, K., Silverman, J. D., Group, CMMID COVID-19 Working, Consortium, COVID-19 Genomics UK (COG-UK), Diaz-Ordaz, K., Keogh, R., Eggo, R. M., Funk, S., Jit, M., Atkins, K. E. and others. (2021). Estimated transmissibility and impact of sars-cov-2 lineage b.1.1.7 in england. Science 372(6538).

Istituto Superiore di Sanitá. (March 2023). Monitoraggio delle varianti del virus SARS-CoV-2 di interesse in sanitá pubblica in Italia. https://www.epicentro.iss.it/coronavirus/sars-cov-2-monitoraggio-varianti-indagini-rapide.

Ito, K., Piantham, C. and Nishiura, H. (2022). Relative instantaneous reproduction number of omicron sars-cov-2 variant with respect to the delta variant in denmark. Journal of medical virology 94(5), 2265–2268.

Kermack, W. O. and McKendrick, A. G. (1927). A contribution to the mathematical theory of epidemics. Proceedings of the Royal Society of London. Series A 115(772), 700–721.

Kurtz, T. G. (1970). Solutions of ordinary differential equations as limits of pure jump markov processes. Journal of Applied Probability 7(1), 49–58.

Liu, Y. and Rocklöv, J. (2021). The reproductive number of the Delta variant of SARS-CoV-2 is far higher compared to the ancestral SARS-CoV-2 virus. Journal of Travel Medicine 28(7).

Liu, Y. and Rocklöv, J. (2022). The effective reproductive number of the omicron variant of sars-cov-2 is several times relative to delta. Journal of travel medicine 29(3).

Lyngse, F. P., Mortensen, L. H., Denwood, M. J., Christiansen, L. E., Møller, C. H., Skov, R. L., Spiess, K., Fomsgaard, A., Lassaunière, R., Rasmussen, M., Stegger, M., Nielsen, C., Sieber, R. N., Cohen, A. S., Møller, F. T., Overvad, M., Mølbak, K., Krause, T. G. and others. (September 2022). Household transmission of the sars-cov-2 omicron variant in denmark. Nature Communications 13(5573).

Presidenza del Consiglio dei Ministri - Dipartimento della Protezione Civile. (2020-2023). https://github.com/pcm-dpc/COVID-19.

R Core Team. (2022). R: A Language and Environment for Statistical Computing. R Foundation for Statistical Computing, Vienna, Austria.

Scalia Tomba, G. (1990). On the asymptotic final size distribution of epidemics in heterogeneous populations. In: Gabriel, J. P., Lefevre, C. and Picard, P. (editors), Stochastic processes in epidemic theory, Lecture Notes in Biomathematics. Springer verlag, pp. 189–195.

Staten Serum Institut. (7th January 2022). Status of the SARS-CoV-2 variant Omicron in Denmark. https://files.ssi.dk/covid19/omikron/statusrapport/rapport-omikronvarianten-07012022-27nk.

UK Health Security Agency. (December 2021). Omicron daily overview: cumulative data. https://assets.publishing.service.gov.uk/government/uploads/system/uploads/attachment_data/file/1044502/sgtf_totalepicurve_2021-12-30.csv/preview.

Volz, E., Mishra, S., Chand, M., Barrett, J. C., Johnson, R., Geidelberg, L., Hinsley, W. R., Laydon, D. J., Dabrera, G., O’Toole, A., Amato, R., Ragonnet-Cronin, M., Harrison, I., Jackson, B., Ariani, C. V., Boyd, O., Loman, N. J., Mc-Crone, J. T., Gonçalves, S., Jorgensen, D., Myers, R., Hill, V., Jackson, D. K., Gaythorpe, K., Groves, N., Sillitoe, J., Kwiatkowski, D. P., consortium, The COVID-19 Genomics UK (COG-UK), Flaxman, S., Ratmann, O., Bhatt, S., Hopkins, S., Gandy, A., Rambaut, A. and others. (2021). Assessing transmissibility of sarscov-2 lineage b.1.1.7 in england. Nature 593(7858), 266–269.

