## Supplementary material for "Estimating the contagiousness ratio between two viral strains"

*Submitted for publication*

### 1. Simulation from the deterministic model

In this section, we present the results obtained on a dataset, simulated with the deterministic discrete-time SIR model described in the paper. The simulated dataset has as true values of the parameters $k = 2$ and $i_0^2 = 20$. We consider a situation with $n = 30$ days, in which we have the information on the couple $(z_j, n_j)$ at 5 different days. The coefficient $\mu$ is set to 1/3 and the total population size is $N = 10^6$. On this dataset, only the deterministic method is tested, and the results obtained are shown in Table 1 and in Figure 1.

| Parameter | Estimate | 95% profile C. I. | 95% C.I. parametric bootstrap |
|---|---|---|---|
| $k$ | 1.95 | $[1.78, 2.16]$ | $[1.79, 2.19]$ |
| $i_0^2$ | 27.9 | $[9.34, 71.69]$ | $[10, 67.3]$ |

Table 1. Results on the deterministic simulated dataset (true values $k = 2$, $i_0^2 = 20$).

*To whom correspondence should be addressed.

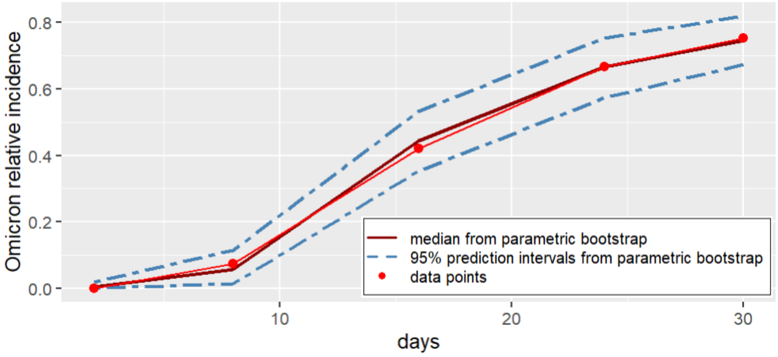


Fig. 1. Estimated relative incidence curve on the deterministic dataset.

### 2. Simulation from the stochastic model

Concerning the stochastic model, two datasets have been simulated, based on the stochastic discrete-time SIR model with Tau-Leap increments described in the paper. The two datasets differ primarily in the values imposed for parameters $k$ and $I_0^2$, and in some other minor aspects. Both datasets have a total population size of $N = 8 * 10^6$. The first dataset has $\mu = 1/3$ and it is simulated for $n = 29$ days in which five data points $(z_j, n_j)$ are known, and the true parameters values are $k = 5$ and $I_0^2 = 2$. The second dataset has $\mu = 1/10$ and it is simulated for $n = 46$ days in which five data points $(z_j, n_j)$ are known, and the true parameters values are $k = 3$ and $I_0^2 = 32$.

#### 2.1 *Estimation with the deterministic method*

The results obtained with the deterministic method on the first dataset are shown in Table 2 and in Figure 2, while those obtained on the second dataset are shown in Table 3 and in Figure 3.

| Parameter | Estimate | 95% profile C. I. | 95% C.I. parametric bootstrap |
|---|---|---|---|
| $k$ | 5.52 | $[4.65, 5.72]$ | $[4.69, 6.73]$ |
| $I_0^2$ | 0.98 | $[0.1, 7.05]$ | $[0.09, 6.14]$ |

Table 2. Results on the first stochastic dataset (true values $k = 5$, $I_0^2 = 2$), deterministic method.

| Parameter | Estimate | 95% profile C. I. | 95% C.I. parametric bootstrap |
|---|---|---|---|
| $k$ | 3.14 | $[2.89, 3.39]$ | $[2.91, 3.45]$ |
| $I_0^2$ | 27.3 | $[24.11, 35.87]$ | $[20.1, 33]$ |

Table 3. Results on the second stochastic dataset (true values $k = 3$, $I_0^2 = 32$), deterministic method.

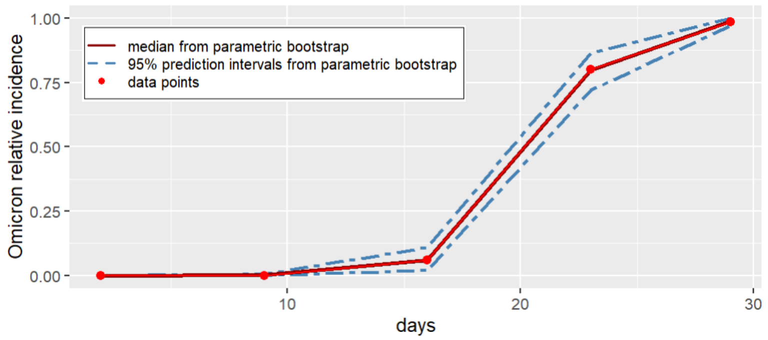


Fig. 2. Estimated relative incidence curve on the first stochastic dataset, deterministic method.

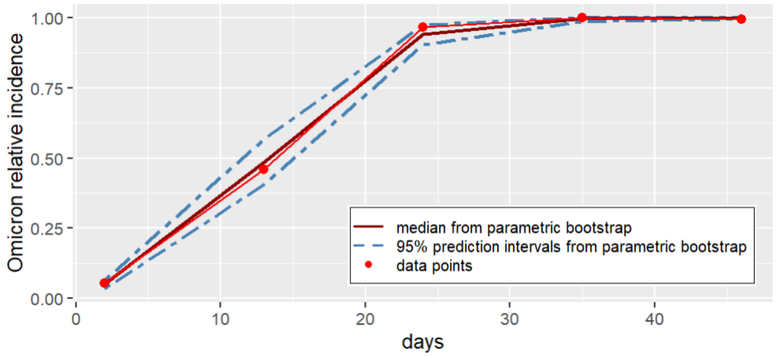


Fig. 3. Estimated relative incidence curve on the second stochastic dataset, deterministic method.

| Parameter | Posterior Median | 95% C. I. |
|---|---|---|
| $k$ | 5.4 | $[4.39, 6.3]$ |
| $I_0^2$ | 2 | $[1, 17]$ |

Table 4. Results on the first stochastic dataset (true values $k = 5$, $I_0^2 = 2$), stochastic method.

| Parameter | Posterior Median | 95% C. I. |
|---|---|---|
| $k$ | 2.95 | $[2.54, 3.37]$ |
| $I_0^2$ | 43 | $[23, 74]$ |

Table 5. Results on the second stochastic dataset (true values $k = 3$, $I_0^2 = 32$), stochastic method.

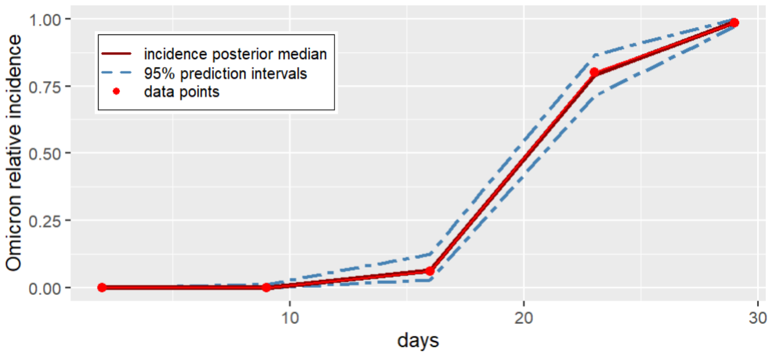


Fig. 4. Estimated relative incidence curve on the first stochastic dataset, stochastic method.

#### 2.2 *Estimation with the stochastic method*

The results obtained with the stochastic method on the first simulated dataset are shown in Table 4 and Figure 4. The results obtained on the second simulated dataset are instead displayed in Table 5 and in Figure 5.

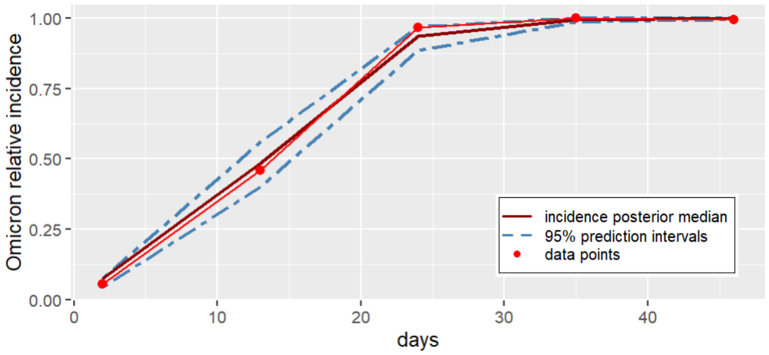


Fig. 5. Estimated relative incidence curve on the second stochastic dataset, stochastic method.
